# Lower-Energy Selective Laser Trabeculoplasty: A Titrated Energy-Pigmentation (TAPE) Approach to Predicting Efficacy and Durability

**DOI:** 10.64898/2026.04.20.26345285

**Authors:** Larry Koval

## Abstract

**Objective:** To evaluate outcomes of lower-energy selective laser trabeculoplasty (SLT) using a titrated energy–pigmentation (TAPE) construct.

**Methods:** This retrospective study analyzed de-identified clinical data of 62 eyes undergoing lower-energy SLT as part of routine care. Laser energy was titrated relative to trabecular meshwork (TM) pigmentation grade and quantified as the product of the two (TAPE score). For example, grade 2 pigmentation × 40 mJ total energy yields a TAPE score of 80. The primary outcome was intraocular pressure (IOP) at 2 months, with adjustment for baseline maximum IOP (Tmax) using analysis of covariance (ANCOVA). Durability was assessed using Kaplan–Meier survival analysis and Cox proportional hazards modeling.

**Results:** After adjustment for baseline Tmax, higher TAPE scores (≥70) were associated with lower 2-month IOP (*p* < 0.01) and greater likelihood of achieving ≥20% IOP reduction. Over longer follow-up, higher TAPE scores were associated with fewer treatment escalation events and improved survival free of additional therapy. Low-grade anterior chamber inflammation was common, transient, and self-limited. At 3 years, 85% of eyes in the high-TAPE group remained drop-free, accounting for retreatment where applicable. No clinically significant IOP spikes or sight-threatening adverse events were observed.

**Conclusions:** Higher energy–pigmentation (TAPE) scores were associated with improved short-term IOP reduction and greater durability following lower-energy SLT. These findings are hypothesis-generating and suggest that TM pigmentation-adjusted energy delivery may enhance biologic engagement of the trabecular outflow pathway and support prospective evaluation of individualized SLT dosing strategies.

## Introduction

Glaucoma is a chronic neurodegenerative disease characterized by progressive optic neuropathy and loss of retinal ganglion cells. It is the most common cause of irreversible blindness in the world and is projected to affect an increasing number of individuals over the coming decades, posing significant societal burden.^1,2^ While multifactorial in pathogenesis and management, the only evidence based validated treatment is reduction of intraocular pressure.^3,4^ Historically, first-line therapy of glaucoma has relied upon topical hypotensive medications. However, eye drop medication may lead to ocular surface toxicity, systemic side effects, and patient daily treatment burden.^5-7^ Lack of adherence to dosing schedules has been well documented and often leads to progression of disease.^8-12^

These challenges have increased the interest in drop-free approaches that provide more consistent IOP control with less daily treatment demands for patients.

Selective Laser Trabeculoplasty (SLT) became available in the United States in 2001. It uses a standard frequency-doubled 532-nm Nd:YAG laser, targeting and absorbed by pigmented cells in the TM. Standard SLT protocols generally apply 100 treatment spots over 360 degrees, with total treatment energy commonly near 90 mJ, and rely upon cavitation bubble formation for titration.^13^

Standard energy SLT has been shown to cause some level of ultrastructural damage to the TM including disruption of the cellular beams.^14,15^ Lower-energy SLT has been pursued as a strategy to improve safety while maintaining efficacy^16-18^

In a long-term retrospective study, Gandolfi demonstrated that annual application of low-power SLT of approximately 25 mJ of total energy resulted in superior 10-year drop-free status, compared with standard SLT performed as needed (PRN)^19^ This finding helped culminate the prospective Trial, Clarifying Optimal Application of SLT Therapy (COAST), which compares fixed low-energy of 40mJ repeated annually versus standard SLT PRN.^20^ Early experience in the COAST Trial informed subsequent protocol refinements, leading to revision in 2025 to initiate treatment with standard SLT followed by re-randomization at year one.^21^

The present study explores whether titrating lower-energy SLT according to TM pigmentation may potentially improve outcomes. The titrated energy-pigmentation (TAPE) construct was developed to quantify treatment intensity.

## Methods

### Study Design

This was a retrospective analysis of de-identified clinical data obtained during routine ophthalmic care and included: 62 eyes with mild to moderate primary open angle glaucoma or ocular hypertension that underwent lower-energy SLT between July 2021 and August 2022.

### SLT Technique

Lower-energy settings ranging from 0.3 to 0.7 mJ/spot were assigned during routine care.

### Trabecular Meshwork Pigmentation Grading and TAPE Score

TM pigmentation was graded gonioscopically, grade 1 to 3. Figure 1 graphically represents the grading system utilized. A single global TM pigmentation grade was assigned to each eye, reflecting the predominant level of pigmentation. The TAPE score was calculated by multiplying total delivered energy by the TM pigmentation grade. For example, grade 2 pigmentation × 40 mJ total energy yields a TAPE score of 80.

**Figure 1.**
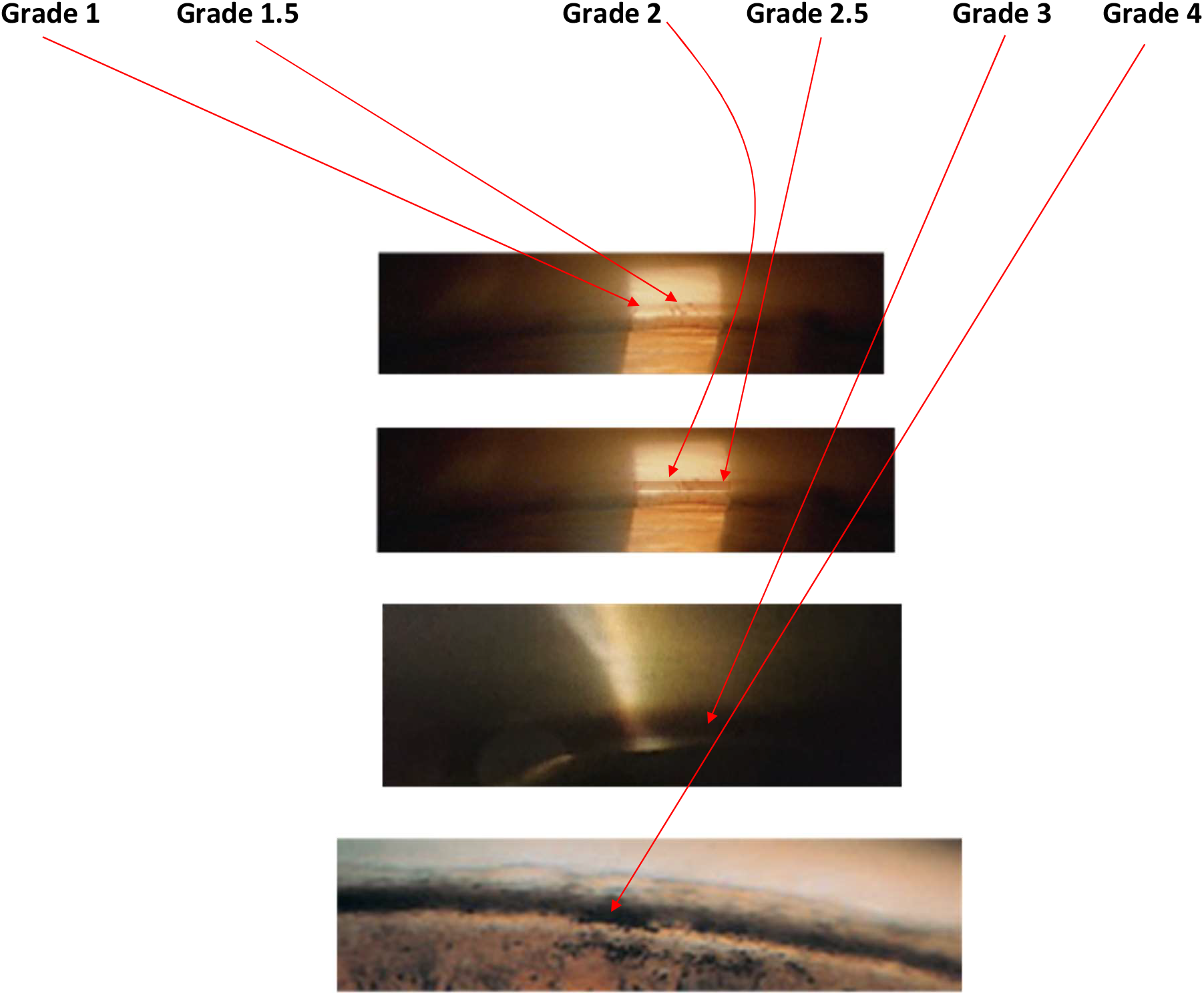
Trabecular Meshwork Pigmentation Grading Scale used.

### Criteria for Escalation of Treatment

IOP ≥ Tmax -2mmHg on two consecutive visits, or progression of disease.

### Outcomes

The primary outcome was IOP at 2 months following SLT. Secondary outcomes included percent IOP reduction, achievement of ≥20% IOP reduction, and durability defined as time to treatment escalation. Baseline IOP was defined as the maximum recorded (Tmax) to reflect real-world clinical decision-making without medication washout. Tmax was used as a covariate to account for inter-eye differences in pretreatment severity, in addition to evidence that peak and fluctuation IOP parameters correlate with glaucoma progression.^3,22-24^.

### Statistical Analysis

The unit of analysis was the eye. Continuous variables are reported as mean ± standard deviation and categorical variables as counts and percentages. Continuous variables were compared using two-tailed t-tests, and categorical variables using standard proportion comparisons.

An ANCOVA model was used to compare 2-month IOP between TAPE groups (Low <70 vs High ≥70) while adjusting for baseline Tmax, with adjusted marginal means estimated at the overall mean Tmax. The relationship between continuous TAPE score and percent IOP reduction was explored descriptively using LOWESS smoothing. Durability was assessed using Kaplan–Meier survival analysis with log-rank testing and Cox proportional hazards modeling. All tests were two-sided with p < 0.05 considered statistically significant.

## Results

Baseline characteristics and laser parameters by TAPE group are shown in Table 1. Eyes in the high-TAPE group demonstrated higher TM pigmentation grades and received greater total delivered energy, by definition, while baseline Tmax and medication use were similar between groups.

**Table 1.**
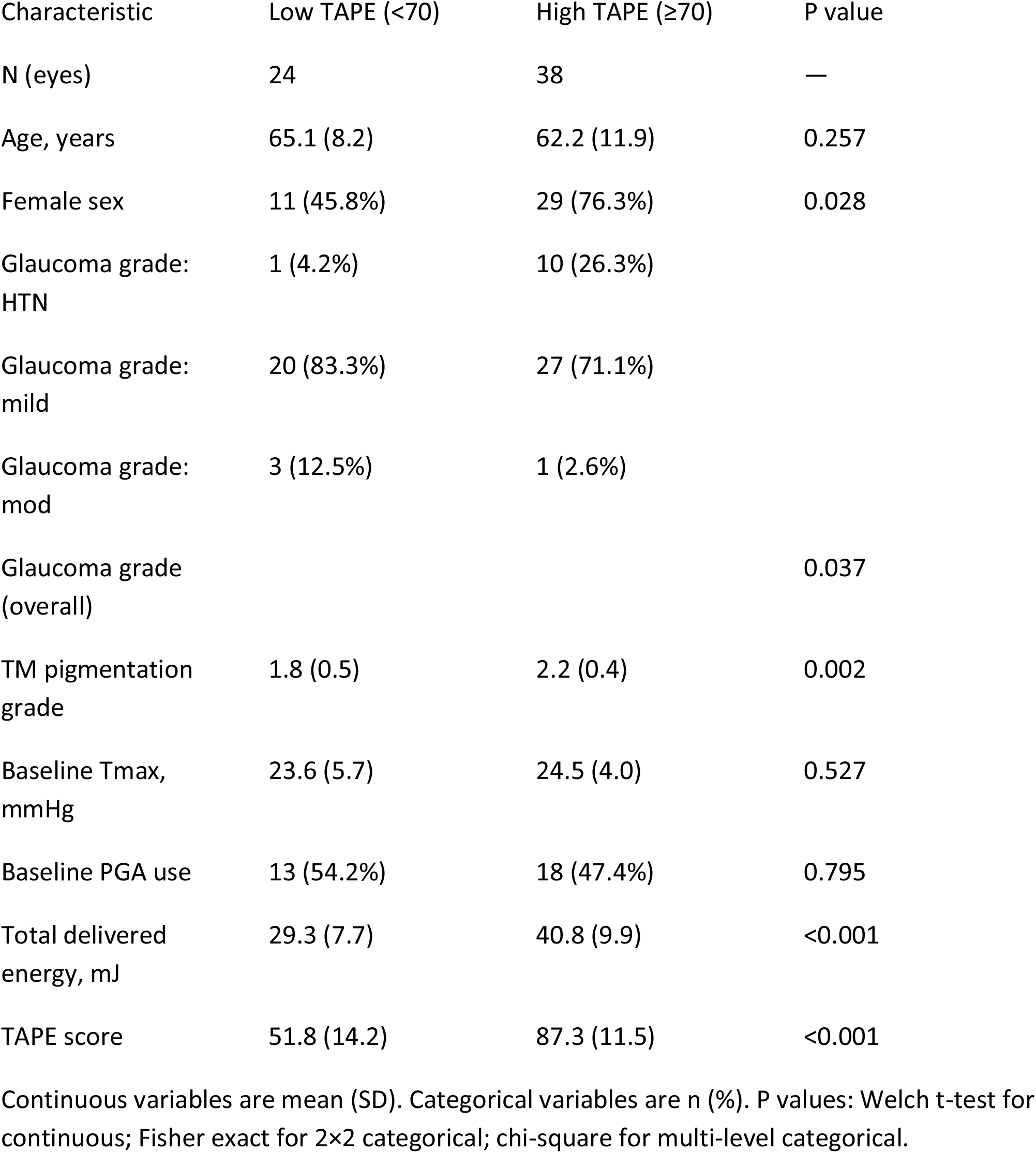
Baseline characteristics and laser parameters by TAPE group (cutoff 70)

Two-month outcomes are summarized in Table 2. After adjustment for baseline Tmax, the high-TAPE group demonstrated lower adjusted mean IOP at 2 months and a greater likelihood of achieving ≥20% IOP reduction.

**Table 2.**
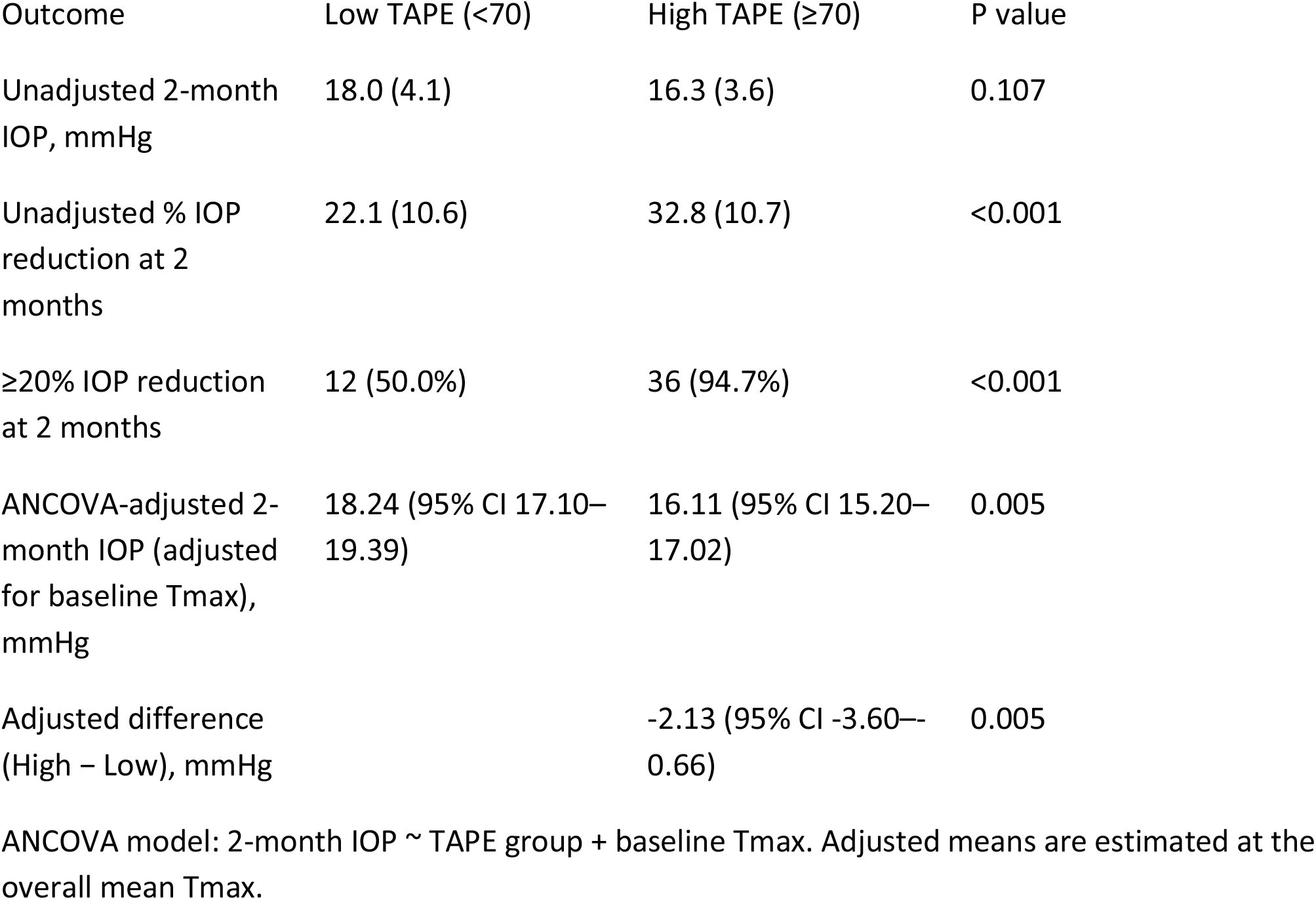
Two-month outcomes by TAPE group, including ANCOVA-adjusted estimates.

The relationship between continuous TAPE score and percent IOP reduction is shown in Figure 2. LOWESS smoothing suggests a positive association between increasing TAPE scores and greater percent IOP reduction; this analysis was descriptive.

**Figure 2.**
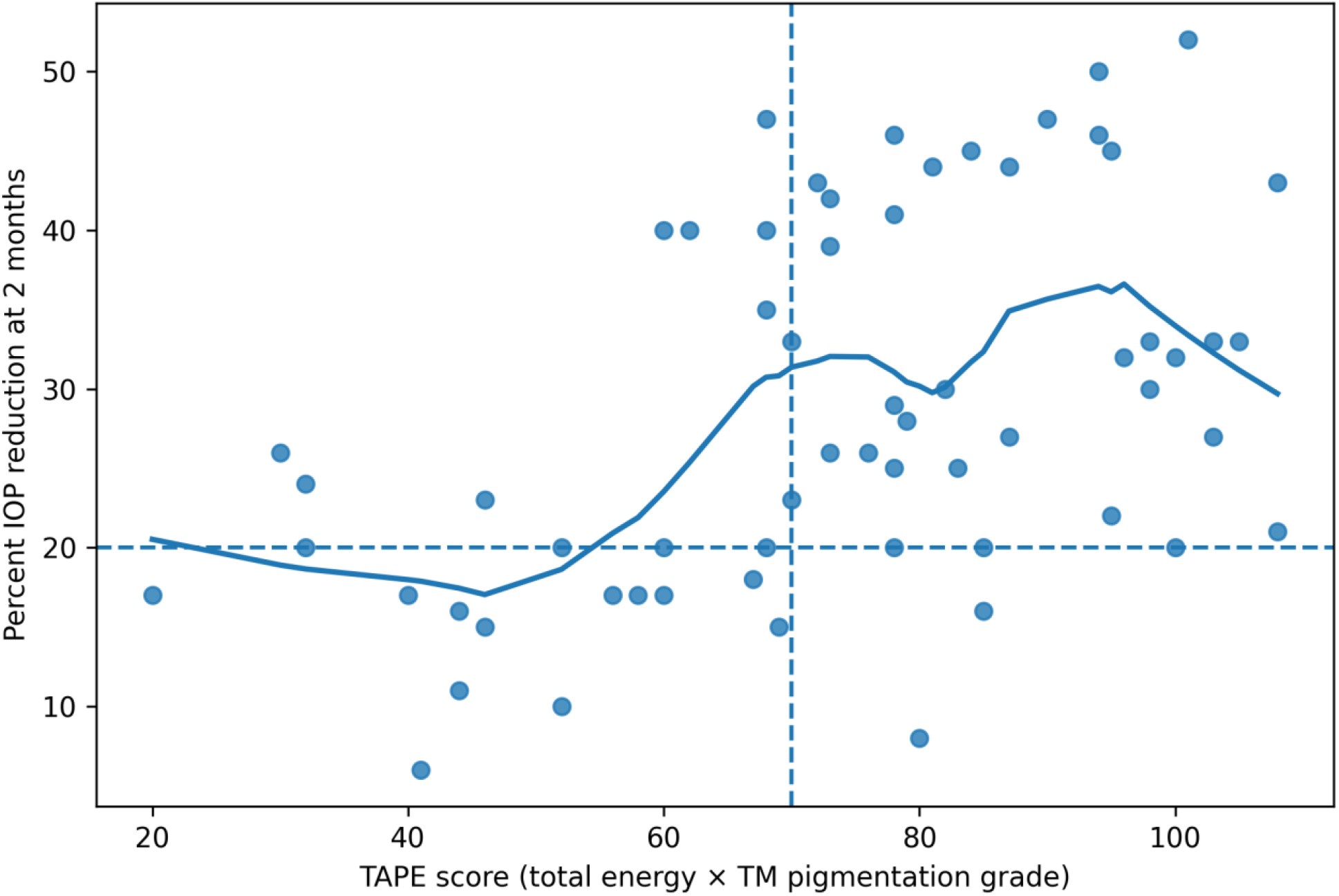
Percent IOP reduction at 2 months by TAPE score (LOWESS) Scatterplot of TAPE score versus percent IOP reduction at 2 months with LOWESS smoothing. Dashed reference lines mark TAPE=70 and 20% IOP reduction.

Durability analyses are shown in Table 3 and Figure 3. Eyes in the high-TAPE group experienced fewer treatment escalation events and improved survival free of escalation over follow-up. Cox proportional hazards modeling demonstrated a reduced hazard of escalation associated with higher TAPE scores.

**Table 3.** Durability and survival analysis (time to treatment escalation) by TAPE group.

| Durability outcome | Low TAPE (<70) | High TAPE (≥70) | P value |
| --- | --- | --- | --- |
| N (eyes) | 24 | 38 | — |
| Escalation events, n (%) | 14 (58.3%) | 7 (18.4%) | — |
| Median time to escalation, months | 24.0 | Not reached | — |
| KM survival at 36 months | 40.1% | 79.8% | — |
| KM survival at 48 months | 40.1% | 79.8% | — |
| Log-rank test (High vs Low) |  |  | 0.001 |
| Cox PH HR (High vs Low) |  | 0.27 (95% CI 0.11–0.68) | 0.005 |

Event defined by Esc Tx=1. Time variable in months. Log-rank p compares Kaplan–Meier curves. Cox PH hazard ratio compares High vs Low TAPE.

**Figure 3.**
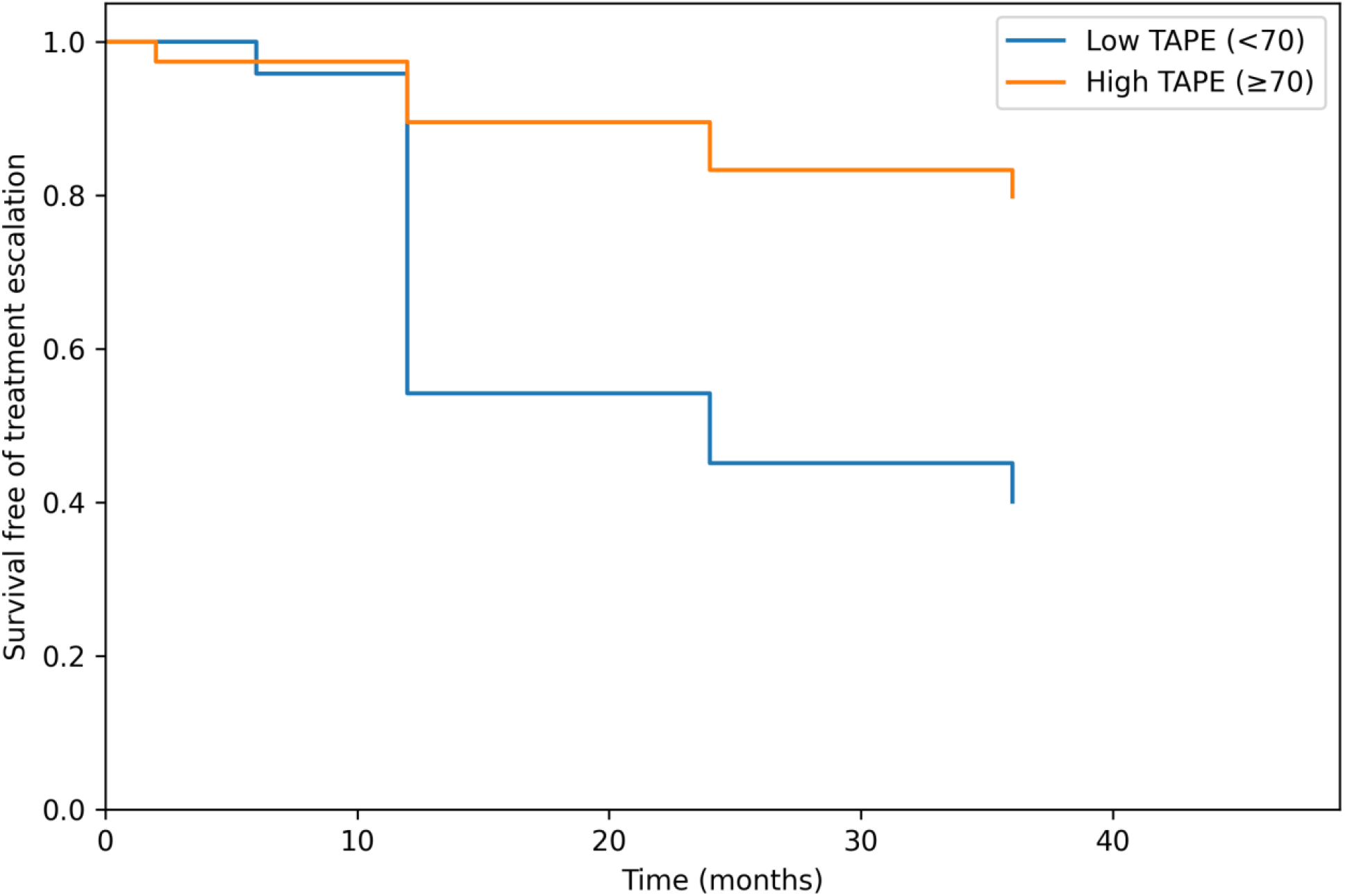
Kaplan–Meier survival free of treatment escalation by TAPE group. Kaplan–Meier curves stratified by TAPE group (cutoff 70). Log-rank p=0.001. Cox PH HR (High vs Low) = 0.27 (95% CI 0.11–0.68).

Low-grade anterior chamber inflammation was commonly observed and rarely required short-term topical corticosteroids. No clinically significant IOP spikes or sight-threatening adverse events occurred.

## Discussion

SLT initiates a biologic cascade involving cytokines and remodeling of the trabecular outflow pathway.^25,26^ Mild anterior chamber inflammation observed in most patients at the 1-hour visit supports a plausible inference that standard SLT and lower-energy SLT have a comparable mechanism of action.

The present findings should be interpreted in the context of lower-energy SLT. Conventional SLT, as originally described, titrates energy relative to TM pigmentation and is performed just below visible microbubble threshold. In contrast, the approach evaluated here consistently operated further below bubble threshold while explicitly quantifying treatment intensity using the TAPE construct. Higher TAPE scores were associated with improved short-term IOP reduction and greater durability, suggesting that biologic engagement of the TM may be achieved across a broader sub-threshold energy range when pigmentation is explicitly considered. While the TAPE metric may not be routinely calculated in clinical practice, it provides a conceptual framework linking energy delivery to TM pigmentation, which may inform a prospective clinical trial to guide individualized SLT dosing.

These findings differ from prior fixed low-energy SLT studies, in which arbitrary energy reduction may have a limited biologic activation. Rather than defining an optimal energy threshold, the present results are hypothesis-generating and support further investigation of pigmentation-adjusted titration strategies.

At baseline, approximately half of eyes were receiving prostaglandin analog therapy, and medication reduction was pursued when clinically appropriate during follow-up. After 3 years, 85% of eyes in the high-TAPE group remained drop-free, accounting for retreatment where applicable.

The LiGHT Trial demonstrated long-term outcomes of standard energy SLT data in 2023.^27^ Approximately 67% of eyes completed the 6-year Trial. 70% were drop-free and 90% of those received 1 to 2 SLT treatments. These data, coupled with adverse event comparisons to eye drop medications, places SLT as perhaps the best initial therapy for primary open angle glaucoma.^27,28^ Lower-energy, and annual versus PRN application continues to be studied, pursuing increased long-term durability—perhaps 10 or more years.^20,21^

Author analysis estimates that lower-energy SLT performed with a high-TAPE score represents 60-70% of standard SLT energy.

Limitations include the retrospective design, the use of 1 clinical site, lack of prospective randomization, and variable follow-up intervals. The TAPE construct has not yet been prospectively validated, and these findings should be interpreted accordingly.

## Data Availability

All data produced in the present study are available upon reasonable request to the authors

## Ethics Statement

This retrospective study utilized clinical data collected during routine patient care. The study protocol was reviewed by an independent institutional review board (Solutions IRB) and determined to be exempt from formal review under 45 CFR 46.104(d)(4) (secondary use of identifiable private information).

*(Exemption ID: 1058; April 17, 2026)*.

## Acknowledgements

The author used a large language model (ChatGPT, OpenAI) for assistance with language editing, organization, and clarification of statistical concepts; all analyses, interpretations, and conclusions were independently performed and verified by the author.

